# Mothers’ knowledge and practices towards oral hygiene of their children aged 5-9 years old: a cross-sectional study in Dhaka

**DOI:** 10.1101/2024.04.05.24305403

**Authors:** Tahazid Tamannur, Sadhan Kumar Das, Arifatun Nesa, Foijun Nahar, Nadia Nowshin, Tasnim Haque Binty, Shafiul Azam Shakil, Shuvojit Kumar Kundu, Md Abu Bakkar Siddik, Shafkat Mahmud Rafsun, Umme Habiba, Zaki Farhana, Hafiza Sultana, Anton Abdulbasah Kamil, Mohammad Meshbahur Rahman

**Affiliations:** Department of Health Education, National Institute of Preventive and Social Medicine, Mohakhali, Dhaka 1212, Bangladesh; Department of Public Health and Hospital Administration, National Institute of Preventive and Social Medicine, Mohakhali, Dhaka 1212, Bangladesh; Directorate General of Health Services, Ministry of Health & Family Welfare, Government of the People’s Republic of Bangladesh; School of the Environment, Nanjing University, Nanjing, China; Dental Speciality Center, Dhaka; BRAC James P Grant School of Public Health, BRAC University, Dhaka; Credit Information Bureau, Bangladesh Bank, Dhaka; Department of Business Administration, Istanbul Gelisim University, Turkey; Department of Biostatistics, National Institute of Preventive and Social Medicine (NIPSOM), Dhaka 1212, Bangladesh

**Keywords:** Knowledge, practice, oral hygiene, mother, Bangladesh

## Abstract

**Background:** Healthy oral hygiene is crucial for overall health and well-being. Parents’ dental care knowledge and practice affect their children’s oral health. Thus, this study assessed the oral hygiene knowledge and practice in mothers of children aged 5-9 years.

**Methods:** This cross-sectional study was conducted from 1 January to 31 December 2022 in Dhaka, Bangladesh. Mothers’ oral hygiene knowledge and practices were assessed through a semi-structured questionnaire. Statistical analysis including the Mann–Whitney U test and Kruskal– Wallis one-way ANOVA test were performed to show average knowledge and practice variations among different socio-demographics of mothers.

**Results:** Out of 400 samples, the mean age of mothers was 30.94±5.15 years where majority were in Muslim faith (97%), housewife (86.8%) and came from nuclear family (68.0%). The prevalence of good knowledge was 41.2%, following 21.5% had moderately average, 18.8% had average and 18.5% mothers had poor knowledge respectively on their children’s oral hygiene. On the other hand, 45.5% mothers had good practice, following 19.5% had average practice, 18.8% had moderately average and 16.2% had poor practice behavior. Mothers’ knowledge levels were significantly (p<0.05) associated with age, education, family size, and monthly income. On the other hand, educational status and income was significantly (p<0.05) associated with mothers’ oral hygiene practices. Non-parametric analysis revealed that the average knowledge level was significantly (p<0.05) higher with respect to higher age group, educational attainment, currently working status, and whose family income was high. On the other hand, oral hygiene practice level was significantly (p<0.05) higher among mother having higher education and higher family income. Mothers’ knowledge was significantly (p<0.05) and positively correlated with the practice behavior obtained by Pearson correlation coefficient.

**Conclusions:** The revealed that the knowledge and practices of mothers directly influence the oral hygiene behavior of children. Mothers with sound knowledge tend to exhibit positive practices concerning their children’s oral hygiene. These findings underscore the importance of taking necessary actions to enhance both the knowledge and practices related to oral hygiene among mothers, thereby ensuring the well-being of their children.

## Introduction

According to the World Health Organization (WHO), dental caries, periodontal disease, tooth loss, mouth cancer, oro-dental trauma, Noma, and congenital defects including cleft lip and palate are classified as oral diseases. Oral health (OH) issues are very prevalent in low-income nations owing to poor socio-educational-economic circumstances (1,2). In terms of general health and well-being, there is a significant connection between OH and overall health (3). It impacts individuals’ capacity to do tasks, communicate, and develop. It also impacts one’s capacity to engage in social interactions. Thus, it has an impact on both the physical and psychological aspects of an individual (4). Most common OH problems and conditions can be readily avoided by establishing suitable oral hygiene routines, such as twice-daily brushing with the best toothbrush, using fluoride-containing toothpaste, and employing the proper brushing technique (5). Other preventive measures include eating a balanced diet low in free sugar, going to the dentist regularly for exams, and getting treatment for illnesses when they are still in the early stages (6). It can be minimized by practicing good oral hygiene habits, such as brushing and flossing teeth and visiting the dentist frequently (7).

Worldwide, over 2 billion individuals have dental caries in their permanent teeth, while 514 million children suffer from dental caries in their primary teeth (8). Early childhood caries (ECC) in children have been linked mostly to poor dental hygiene. Infants and toddlers with significant plaque accumulation were more likely to experience severe ECC and caries from birth to toddlerhood (9). ECC is primarily caused by several causes, including excessive sugar intake, poor dental hygiene, inadequate fluoride exposure, and enamel abnormalities (10). So, the development of caries and the acquisition of infection are significantly influenced by diet and feeding habits.

As parents are the major caregivers, their involvement is crucial in the maintenance and development of excellent oral health of a child, such as teaching healthy eating and drinking habits (11). In addition, several other factors impact the dental health of children, including the mother’s level of education, the mother’s work situation, and her understanding of oral hygiene (12). That means, the adoption of healthy oral health practices in children is influenced by parents’ particularly mothers’ oral health knowledge, attitudes, and awareness (13). An Indian study found that the oral hygiene quality of children at 12 years old was shown to be significantly influenced by their mothers’ oral hygiene knowledge (14). Children with high rates of dental caries and low rates of fillings were found to have parents with inadequate oral health literacy, according to another research (15). As a result, it is essential for parents, and particularly mothers, to have awareness about oral health. Scholars argued that mothers’ knowledge about oral health and the consequence of inadequate dental hygiene has a beneficial impact on their children’s dental well-being and adherence to dental care practices (16,17).

In research on dental caries awareness among parents in Pakistan, found that there were low levels of knowledge of oral hygiene standards (18). A study conducted in India on the oral health status of 3–6-year-old children and their mother’s oral health-related knowledge, attitude, and practices found most mothers had a medium level of knowledge, an average level of attitude, and a high level of practices regarding oral health (19). Another study in Malaysia on parental knowledge and practices in preschool children’s oral healthcare found that the majority had good knowledge (20).

Numerous studies have been conducted globally regarding parents’ or mothers’ oral hygiene knowledge and practices. However, the current study on the knowledge and practice of oral hygiene among parents, particularly mothers, in Bangladesh is insufficient. There is a lack of research investigating the extent to which moms are aware of and follow oral hygiene practices. Hence, this study aimed to fill this research gap. The objective of this research was to assess the level of knowledge and practice of oral hygiene habits among mothers, while also identifying the factors that influence these practices.

## Methods

### Ethical consent and permission for data collection

The permission for this study was approved by the institutional review board of the National Institute of Preventive and Social Medicine (NIPSOM), Bangladesh (Ref No: NIPSOM/IRB/2017/09). Shaheed Suhrawardy Medical College Hospital and Dhaka Dental College Hospital provided the necessary documentation. Participants received an overview of the study’s goals and those who consented were eventually included.

### Study setting and participants

This cross-sectional study was conducted from 1 January to 31 December 2022 in two tertiary-level hospitals named Shaheed Suhrawardy Medical College Hospital and Dhaka Dental College Hospital in Dhaka City. Mothers of children aged between 5 to 9 years were the study participants. A questionnaire comprised of the socio-demographic variables and knowledge and practice of oral hygiene was provided to fill.

### Sampling technique and sample size

A convenient sampling technique was followed for this study. The sample size was calculated using the equation below.

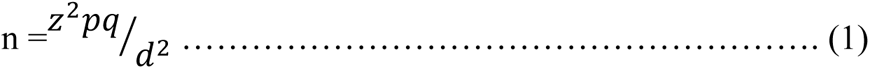

the sample size for the mother’s knowledge when p=0.58 (21) was

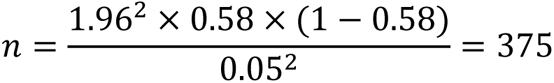

Similarly, the sample size for mother’s practice level when p=0.57 (21) was

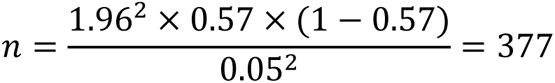

Therefore, we initially chose a maximum of 377 as the required sample size. Considering a maximum 5% non-response rate (based on pre-testing), we rounded up this figure and selected 400 as the approximate sample size in the study.

### Selection criteria

The inclusion criteria for this study were (i) mothers with Bangladeshi nationality who were living in Dhaka for at least one year; (ii) mothers of children aged 5-9 years; and (iii) mothers who provided consent and agreed to participate in the study. The exclusion criteria for the study were (i) mothers who were not Bangladeshi but currently living in Dhaka; (ii) mothers of children aged more than 10 and less than 5; (iii) mothers aged less than 21 and more than 48 years old.

### Outcome measures

#### Socio-demographic Variables

Respondents’ socio-demographic variables such as age (21–48), religion (Muslim, Non-Muslim), educational status (up to primary, secondary, higher secondary, graduation or above), occupational status (housewife, working), family type (nuclear, joint), family size (less than 5 persons, equal or greater than 5 persons), and monthly family income (≤ 20000 BDT, 21000-40000 BDT, ≥ 41000 BDT) were independent.

#### Measurement of knowledge and practice score

The study used 15 variables to assess mothers’ knowledge and 13 to assess practice-related oral hygiene. The summation scoring technique was used in computation, and their descriptive statistics, including percentiles, were observed. According to the percentile approach, the knowledge was classified into four levels-poor knowledge (<25.0% percentile cut-point: ≤9.999); moderately average (25.0%-49.0% percentile, cut point:10.0-11.99); average knowledge (50.0%-74.0% percentile, cut point: 12.0-12.99); and good knowledge (≥75.0% percentile, cut point: ≥13.0) (22). The practice was also classified into four levels-poor practice (<25.0% percentile cut-point: ≤ 4.99); moderately average practice (25.0%-49.0% percentile, cut point: 5.0-5.999); average practice (50.0%-74.0% percentile, cut point: 6.0-6.99) and good practice (≥ 75.0% percentile, cut point: ≥ 7.0). For all cases, the cut points were statistically evident (23).

#### Statistical analysis

Descriptive statistics were performed to present participants’ socio-demographic characteristics and mean knowledge and practice scores. Since both knowledge and practice scores did not follow normality, we performed the Mann–Whitney U test and Kruskal–Wallis one-way ANOVA test to show the mean knowledge and practice variation between two (e.g., housewife versus working mother) and more than two (e.g.: different age groups) groups respectively. P-value was observed for all the cases at 5% level and 95% was considered as the confidence interval (24).

## Results

### Socio-demographic characteristics of the respondents

Table 1 showed the socio-demographics portion of the study. More than half (52.2%) of the respondents were within the age group 21-30 years followed by 44.0% in between the age group 31-40 years. The highest (39.2%) respondents were at the secondary level. Most were Muslims (97%) and housewives (86.8%). Most respondents (39.3%) had a monthly family income of 21000-40000 taka per month, followed by 13.3% were working mothers (Table 1).

**Table 1.**
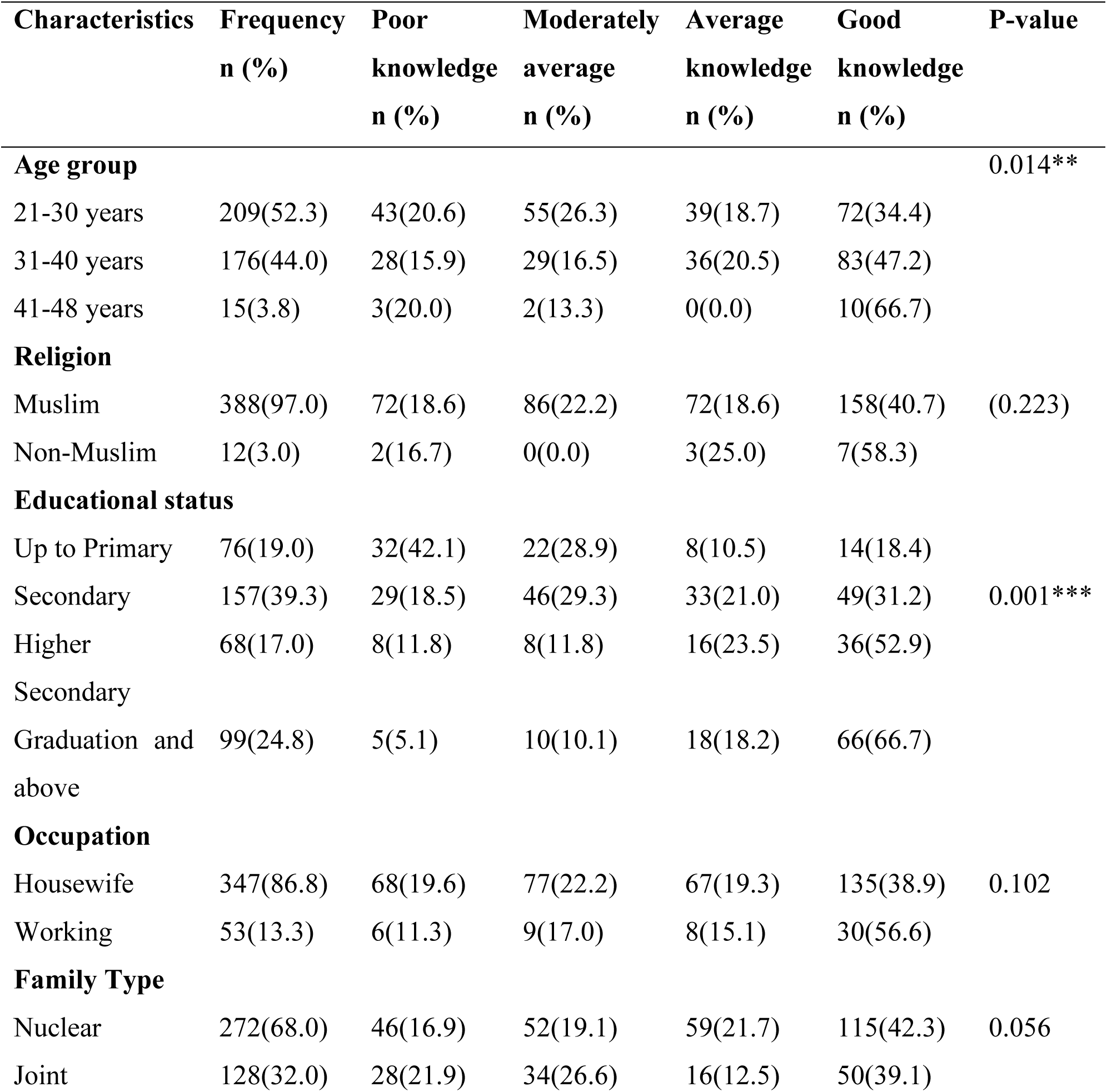
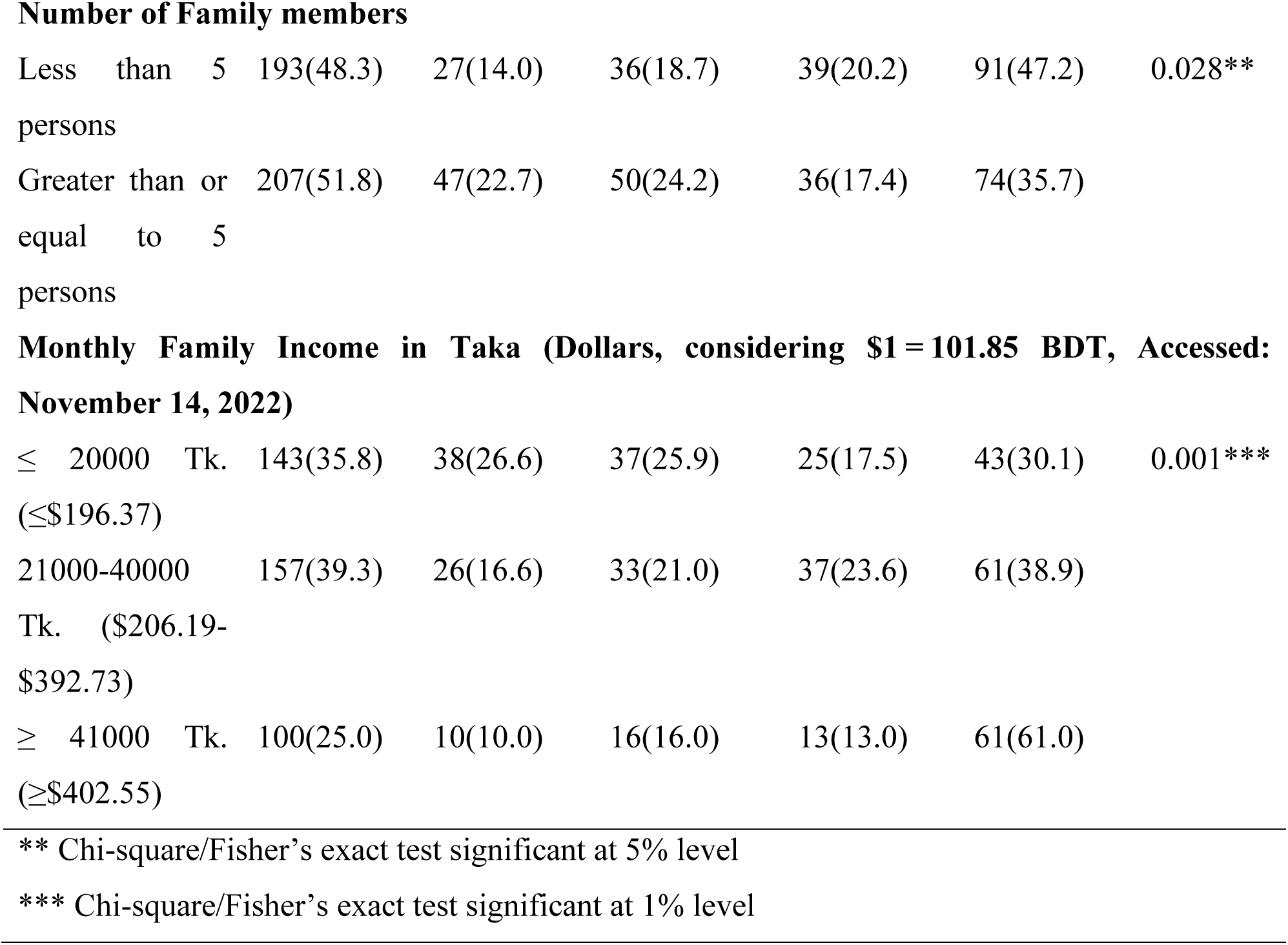
Association of Mothers’ knowledge with socio-demographic characteristics.

### Mothers’ individual knowledge and practice regarding children’s oral hygiene

Out of 400 mothers, more than 90% knew the importance of brushing teeth while 82.3% and 80.8% of them knew the recommended frequency and appropriate time for brushing teeth. Surprisingly, only 29.5% and 38.5% knew the duration for brushing teeth and fluoride can give protection against caries. However, most of the respondents knew about ‘importance of cleaning tongue’ (91.3%), ‘gingival disease common cause of gum bleeding’ (71.5%), ‘brushing and flossing protect against bleeding gum’ (60.8%), ‘yellow coating plaque’ (90.5%), ‘sugary item cause caries’ (96.8%), ‘soft drinks cause caries’ (73.8%), ‘regular brushing protects against caries’ (95.0%) (Supplementary Table S1).

On the other hand, 95.3% of the mothers pointed that their child brushed their teeth regularly, 99.0% of children used a toothbrush, 62.0% changed their toothbrush 3-4 months or if the bristle frayed out, 97.8% used their toothpaste, 77.8% rinsed their mouth after eating. Surprisingly, 44.3% of children brushed their teeth twice daily, 42.0% cleaned their tongues, and 2.8% used floss. Only 12.5% were given sugary items with meals, and 0.3% were taken to dentists every 6 months (Supplementary Table S2).

### Overall knowledge and practice level of the respondents

Fig 1 depicted the level of knowledge and practices of mothers regarding their children’s oral hygiene and their association with mothers’ educational status. Only 41.2% had good knowledge, while 18.5% had poor knowledge (Fig 1a). Similarly, only 45.5% of the mothers had good practice, while 16.2% had poor practice levels (Fig 1b).

**Figure 1.**
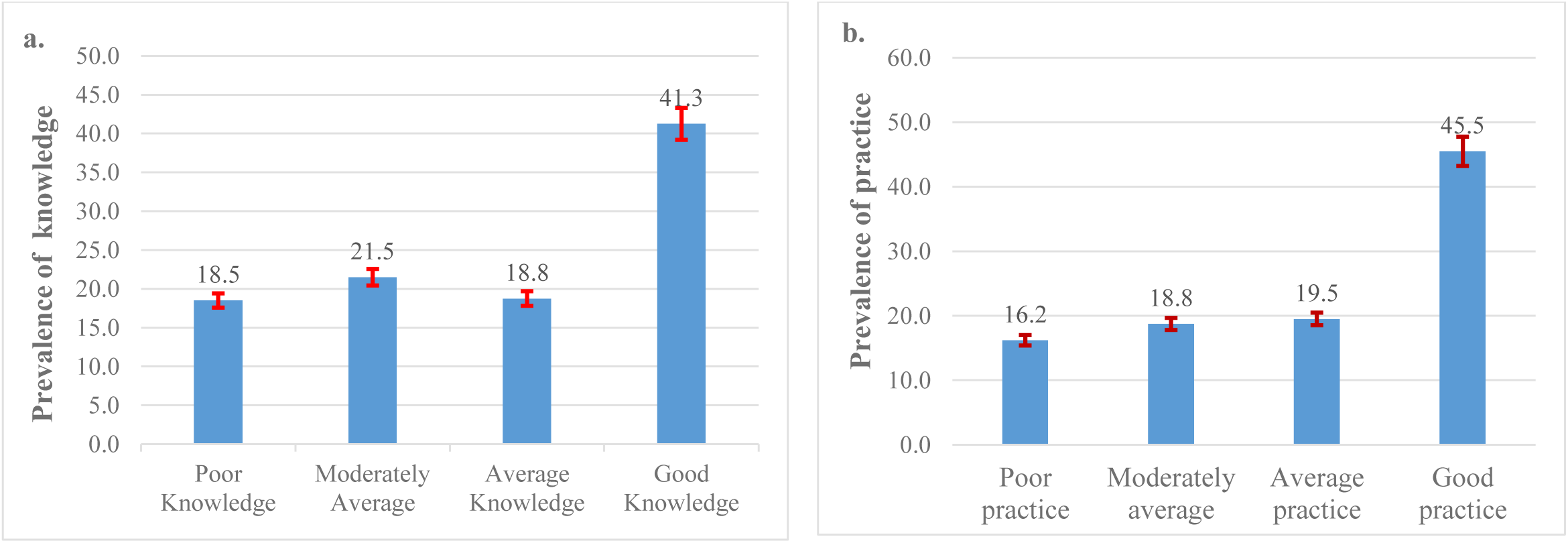
(1.a) Distribution of overall knowledge; (1.b) Distribution of overall practice of mothers.

### Association of sociodemographic characteristics with mothers’ knowledge and practices

In the age group 41-48 years, 66.7% had good knowledge. The association between the age of mothers and knowledge level under null hypothesis H0 (H0: there is no association between the age of the mothers and knowledge level) was significant (p=0.014). Among 388 Muslim mothers’ the majority (40.7%) showed good knowledge and not statistically significant (p=0.223) association was present between knowledge level and religion. In this study, a significant (*p=0.001*) association was also found between educational status and knowledge level (p=0.001); the number of family members and knowledge level (p=0.028); and family income and knowledge level (Table 1). On the other hand, more than half (53.3%) of older-aged, and 44.8% of Muslim mothers had good practices. The majority (66.7%) of graduated mothers had a good practice and educational level was significantly associated with practice level (p=0.002). More than 50.0% of mothers with higher-income families had significantly good practices behaviors (Table 2).

**Table 2.**
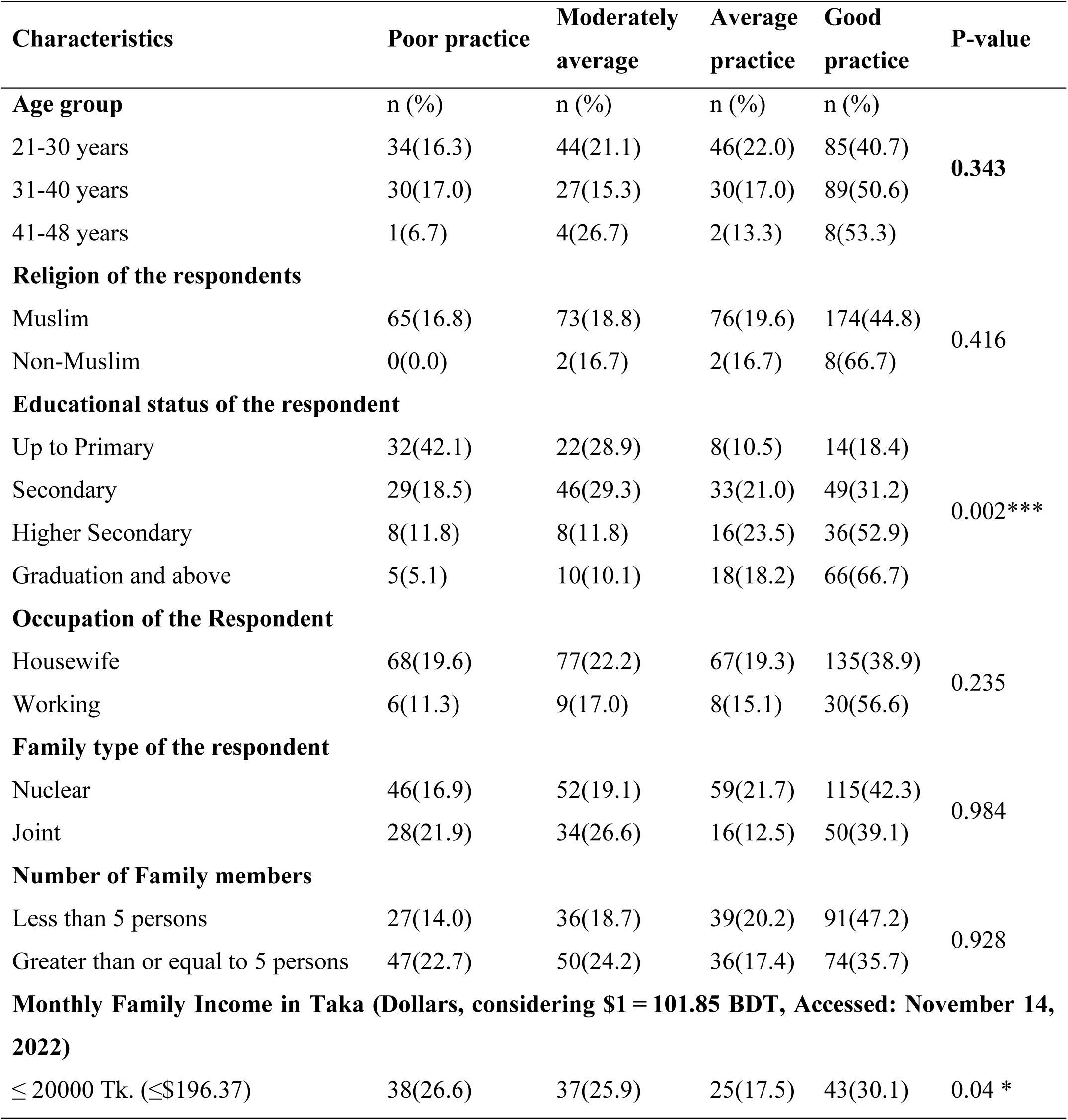
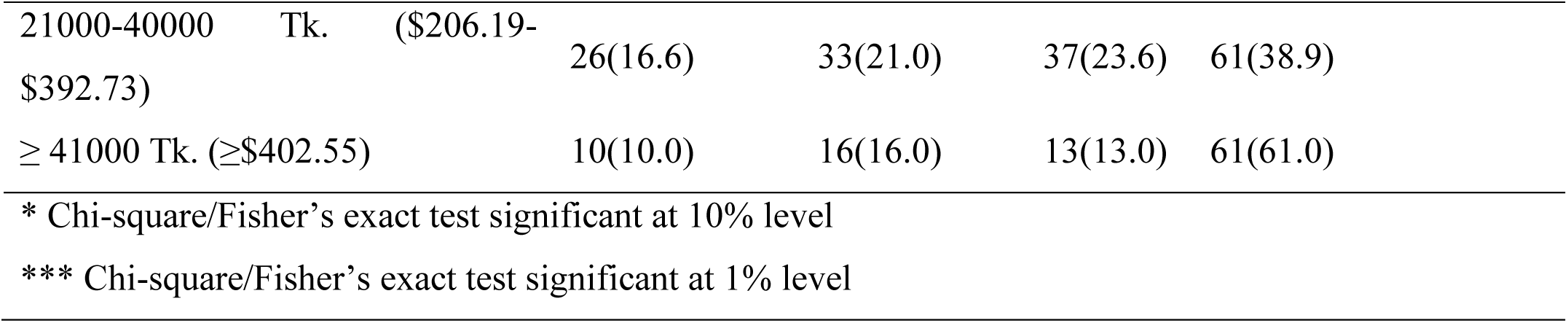
Association of mothers’ practice behavior with socio-demographic characteristics.

| Characteristics | Poor practice | Moderately average | Average practice | Good practice | P-value |
| --- | --- | --- | --- | --- | --- |
| **Age group** | n (%) | n (%) | n (%) | n (%) |  |
| 21-30 years | 34(16.3) | 44(21.1) | 46(22.0) | 85(40.7) | **0.343** |
| 31-40 years | 30(17.0) | 27(15.3) | 30(17.0) | 89(50.6) |  |
| 41-48 years | 1(6.7) | 4(26.7) | 2(13.3) | 8(53.3) |  |
| **Religion of the respondents** |  |  |  |  |  |
| Muslim | 65(16.8) | 73(18.8) | 76(19.6) | 174(44.8) | 0.416 |
| Non-Muslim | 0(0.0) | 2(16.7) | 2(16.7) | 8(66.7) |  |
| **Educational status of the respondent** |  |  |  |  |  |
| Up to Primary | 32(42.1) | 22(28.9) | 8(10.5) | 14(18.4) | 0.002*** |
| Secondary | 29(18.5) | 46(29.3) | 33(21.0) | 49(31.2) |  |
| Higher Secondary | 8(11.8) | 8(11.8) | 16(23.5) | 36(52.9) |  |
| Graduation and above | 5(5.1) | 10(10.1) | 18(18.2) | 66(66.7) |  |
| **Occupation of the Respondent** |  |  |  |  |  |
| Housewife | 68(19.6) | 77(22.2) | 67(19.3) | 135(38.9) | 0.235 |
| Working | 6(11.3) | 9(17.0) | 8(15.1) | 30(56.6) |  |
| **Family type of the respondent** |  |  |  |  |  |
| Nuclear | 46(16.9) | 52(19.1) | 59(21.7) | 115(42.3) | 0.984 |
| Joint | 28(21.9) | 34(26.6) | 16(12.5) | 50(39.1) |  |
| **Number of Family members** |  |  |  |  |  |
| Less than 5 persons | 27(14.0) | 36(18.7) | 39(20.2) | 91(47.2) | 0.928 |
| Greater than or equal to 5 persons | 47(22.7) | 50(24.2) | 36(17.4) | 74(35.7) |  |
| **Monthly Family Income in Taka (Dollars, considering \$1 = 101.85 BDT, Accessed: November 14, 2022)** | | | | | |
| ≤ 20000 Tk. (≤\$196.37) | 38(26.6) | 37(25.9) | 25(17.5) | 43(30.1) | 0.04 * |

|  |  |  |  |  |
| --- | --- | --- | --- | --- |
| 21000-40000 Tk. (\$206.19-\$392.73) | 26(16.6) | 33(21.0) | 37(23.6) | 61(38.9) |
| ≥ 41000 Tk. (≥\$402.55) | 10(10.0) | 16(16.0) | 13(13.0) | 61(61.0) |
| * Chi-square/Fisher's exact test significant at 10% level |  |  |  |  |
| *** Chi-square/Fisher's exact test significant at 1% level |  |  |  |  |

### Mothers’ knowledge and practices variations regarding their children’s oral hygiene

A significant difference in respondents’ knowledge and practice with socio-demographic characteristics was observed (Table 3). Results found that the knowledge was comparatively higher [mean knowledge score among 41 to 48 aged mothers was 12.133 (10.73-13.54)] among older than younger. Similarly, knowledge and practice behavior were significantly (p<0.05) higher among higher educated mothers compared to least educated ones and higher income groups than lower. In addition, working mothers and mothers with small families had significantly higher knowledge (Table 3).

**Table 3.** Knowledge and practice variation of mothers according to sociodemographic characteristics.

| Characteristics | N | Mean knowledge | P-value | Mean practice | P-value |
| --- | --- | --- | --- | --- | --- |
| **Age group** |  |  |  |  |  |
| 21-30 years | 209 | 11.21(10.85-11.58) | 0.01** | 6.13(5.92-6.35) | 0.205 |
| 31-40 years | 176 | 11.93(11.56-12.29) |  | 6.36(6.09-6.62) |  |
| 41-48 years | 15 | 12.13(10.73-13.54) |  | 6.8(5.67-7.93) |  |
| **Religion** |  |  |  |  |  |
| Muslim | 388 | 11.54(11.28-11.80) | 0.216 | 6.24(6.07-6.41) | 0.194 |
| Non-Muslim | 12 | 12.25(10.53-13.97) |  | 6.83(6.08-7.59) |  |
| **Educational status** |  |  |  |  |  |
| Up to Primary | 76 | 9.66(8.95-10.37) | 0.0001*** | 6.01(5.63-6.40) | 0.001*** |
| Secondary | 157 | 11.32(10.97-11.67) |  | 6.01(5.75-6.27) |  |
| Higher Secondary | 68 | 12.26(11.71-12.82) |  | 6.19(5.79-6.59) |  |
| Graduation and above | 99 | 12.93(12.55-13.31) |  | 6.88(6.54-7.22) |  |
| **Occupation** |  |  |  |  |  |
| Housewife | 347 | 11.45(11.17-11.73) | 0.027** | 6.21(6.02-6.39) | 0.12 |
| Working | 53 | 12.30(11.72-12.89) |  | 6.59(6.19-6.98) |  |
| **Family type** |  |  |  |  |  |
| Nuclear | 272 | 11.7(11.39-12.00) | 0.131 | 6.25(6.05-6.45) | 0.878 |
| Joint | 128 | 11.28(10.81-11.75) |  | 6.28(5.98-6.59) |  |
| **Number of family members** |  |  |  |  |  |
| Less than 5 persons | 193 | 11.96(11.6-12.32) | 0.001*** | 6.27(6.03-6.51) | 0.949 |
| ≥5 persons | 207 | 11.19(10.84-11.55) |  | 6.25(6.01-6.48) |  |
| **Monthly income** |  |  |  |  |  |
| Less than 20000 | 143 | 10.92(10.48-11.36) | 0.001*** | 5.96(5.68-6.24) | 0.002*** |
| 21000-40000 | 157 | 11.56(11.17-11.95) |  | 6.20(5.96-6.45) |  |
| Greater than 41000 | 100 | 12.49(12.0-12.98) |  | 6.77(6.40-7.14) |  |

\*\* Significant at 5% level
\*\*\*Significant at 1% level

### Association between mothers’ oral hygiene knowledge and practice level

Figure 2 represented association between mothers’ oral hygiene knowledge and practice level. Over 50% of mothers with higher knowledge had good practices. A statistically significant association was present between knowledge level and practice level (p=0.003).

**Figure 2.**
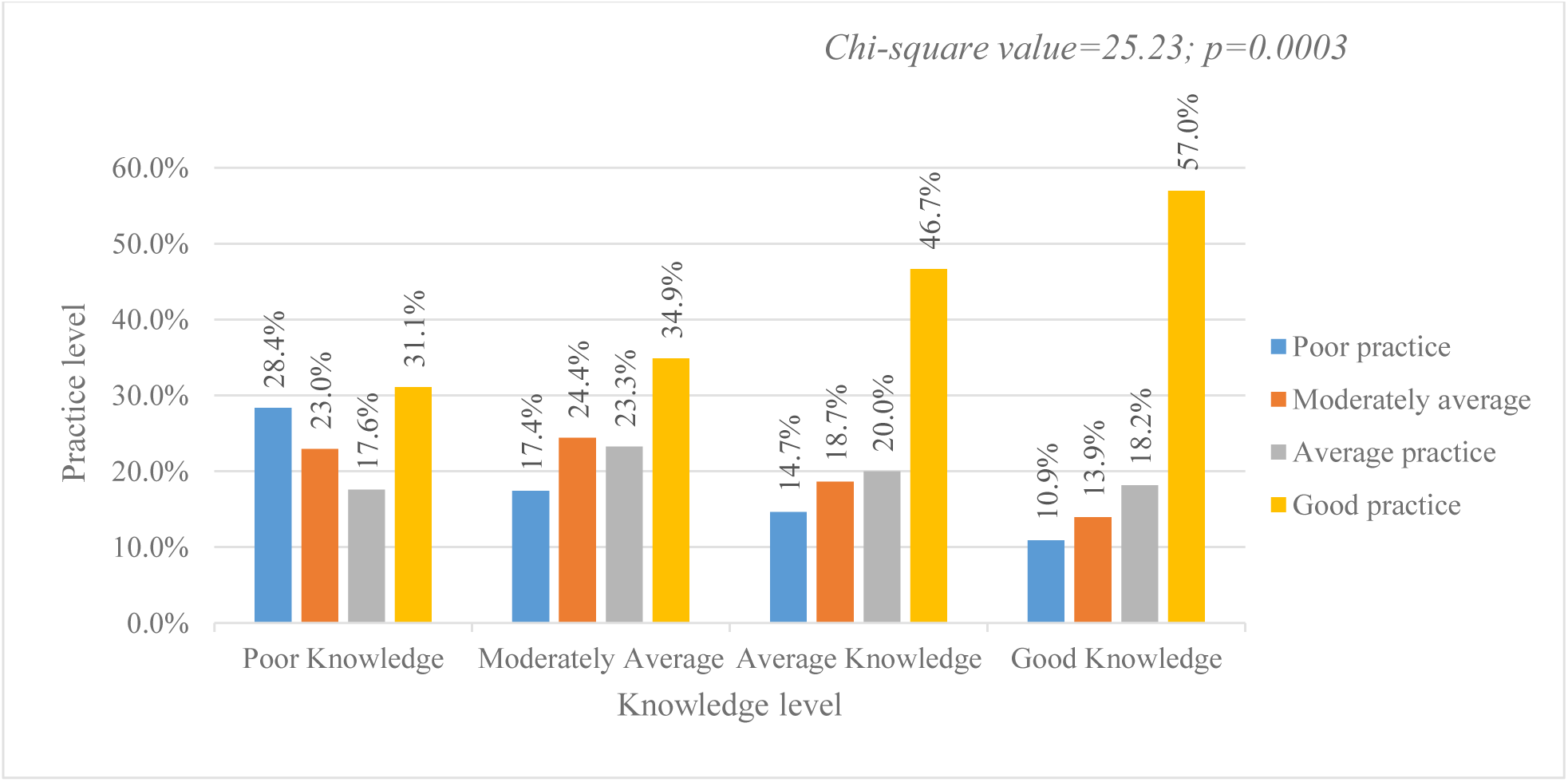
Association between mothers’ knowledge level and practice level

## Discussion

Oral health is an integral component of overall health, and its significance in our everyday lives is paramount. This study intended to evaluate parents’ knowledge and practices regarding their children’s oral hygiene. An increase in knowledge level was observed in older adults, higher education levels, working mothers, and higher family income respondents. Similarly, good practices were associated with education level and economic status.

Parents are generally conscious about the importance of their children’s tooth brushing, and almost all mothers found knowledgeable regarding this. Most mothers know brushing twice a day is the standard recommendation, which is comparable to the existing study (25,26). In our study, many mothers thought gingival disease was the most common cause of gum bleeding, and more than half thought brushing, and flossing can protect against bleeding gum that is comparable to the previous literature (27). The majority mothers knew about the bacterial transmission by using the same utensils which is comparable to the existing studies (28).

American Dental Association (ADA) recommends that a toothbrush should be changed in 3-4 months or sooner if the bristle frays out (29). In this study, more than half of the mothers agreed with the statement and they changed their child’s brush between 3-4 months or if the bristle frayed out. Almost all mothers used toothpaste when cleaning their child’s teeth cleaning. According to a study, most participants used a toothbrush and a fluoride-containing paste to brush their children’s teeth, but only a small minority of moms in the study were aware that they did so (26). It has been reported that tongue scraping and brushing act as an excellent method for lowering plaque levels in the mouth (4). The ADA recommends using dental floss once every day to effectively remove microbial plaque and avoid gingivitis. Almost all mothers said they did not take their child to the dentist every 6 months for regular checkups.

The findings of the present study indicated that almost half of mothers had good knowledge regarding their child’s oral hygiene which is consistent with the study of Bakar and Mamat (20). But Shalini et al. found that 14.5% had excellent knowledge (30) and Abduljalil and Abuaffan, in their study showed that mothers had good knowledge (31). Findings also revealed that a maximum of the mothers had good oral hygiene practice, but Krishnan et al. found that 13.7% of the mothers had good practice (29). A significant association was present between knowledge level and age of the mothers in contrast with the study of Al-Jaber et al. (32). Likewise educational status and monthly family income are significantly associated with knowledge level. Similarly significant association between education and knowledge was found in the study of Olatosi and other authors (33). On the other hand, Moslemi et al. found a significant association between educational status, income, and occupation with knowledge regarding oral hygiene (34). However, in the present study, there is no association between occupation and knowledge level. More educated parents are more concerned with keeping their children’s healthy dentition and have a more positive outlook on their children’s oral health. In the current study, the association between practice and educational status was statistically significant which is like the study of Alzaidi et al. (23). Mothers in the age group 41 to 48 years showed significantly higher knowledge compared to other age groups. However, the difference in practice scores were not statistically significant among age groups which is comparable to a study done in India (35). Mothers with graduation and above degrees scored higher mean knowledge and practice scores compared with others. It was observed that the working mothers had significantly higher mean knowledge. The findings were comparable to a study done in India (35). Significant differences in knowledge scores as well as practice scores among different income groups were also observed.

The people of Bangladesh have been facing many communicable and non-communicable diseases in different times (36–46) where oral health problem is the neglected one due to low-economic and social status (2,37). The objective of this study was to identify the variables that impact oral hygiene habits among mothers and to evaluate their level of awareness and compliance with oral hygiene practices. The primary merit of this study lies in its research results. We identified the variables that influence individuals’ understanding and behaviors related to oral hygiene. Although the study’s cross-sectional pattern and sample size are limited, it will nonetheless provide valuable insights for policymakers to make informed decisions. To get a more extensive perspective, more studies might be undertaken using a larger and more complete sample size.

The results of the study suggest including the implementation of oral health education programs, community outreach activities, and the integration of oral health education into primary healthcare. It is essential to prioritize the provision of dental treatments that are easily accessible, implement programs in schools, and carry out media campaigns. To achieve lasting improvement in mothers’ knowledge and habits about their children’s oral hygiene, it is crucial to promote parental participation in schools, establish monitoring systems, cultivate relationships with non-governmental organizations (NGOs), and provide ongoing funding for research.

## Conclusion

The main purpose of oral hygiene is prevention. This implies that oral health issues like cavities, gum disease, bad breath (halitosis), and other issues can be avoided before they start by taking good care of one’s teeth and gums. Dental hygiene is a critical aspect of a child’s general health and a significant predictor of the standard of living for children. Parental understanding and practices are the main variables that have a direct impact on how children behave and how their dental health is influenced. Many difficulties arise because of improper oral hygiene maintenance knowledge. Children are unable to effectively make their own decisions. Therefore, parents, especially mothers, need to be very careful about their kids’ oral health. The relevant parties must take the required steps to educate women about proper dental hygiene to ensure the safety of themselves and their children.

## Acknowledgments

We acknowledge the Department of Biostatistics, National Institute of Preventive and Social Medicine (NIPSOM) for their technical support during the study. We are also grateful to all participants included in this study.

## Conflict of interest

The authors declared no conflicts of interest exist.

## Funding statement

This study received no specific funds from any agencies or organizations.

## Ethics approval and consent from the participants

This study was approved by the institutional review board of the National Institute of Preventive and Social Medicine (NIPSOM), Bangladesh (Ref No: NIPSOM/IRB/2017/09). Both written and verbal consent were taken before initiating the interview. A brief on the aims and objectives was given to the participants. Participants who agreed were finally included in the study.

old: a cross-sectional study in Dhaka

## Authors contributions

**Conceptualizations-** Mohammad Meshbahur Rahman, Hafiza Sultana, and Tahazid Tamannur

**Formal analysis-** Tahazid Tamannur, Sadhan Kumar Das and Mohammad Meshbahur Rahman

**Investigation-** Tahazid Tamannur, Sadhan Kumar Das, Arifatun Nesa, Arifatun Nesa, Foijun Nahar, Nadia Nowshin, Tasnim Haque Binty, Shafiul Azam Shakil, Shuvojit Kumar Kundu, Md Abu Bakkar Siddik, Shafkat Mahmud Rafsun, Umme Habiba, and Zaki Farhana

**Methodology-** Tahazid Tamannur, Zaki Farhana and Mohammad Meshbahur Rahman

**Project administration-** Tahazid Tamannur, Hafiza Sultana, Anton Abdulbasah Kamil, Mohammad Meshbahur Rahman

**Supervision-** Mohammad Meshbahur Rahman

**Validation-** Tahazid Tamannur, Zaki Farhana and Mohammad Meshbahur Rahman

**Visualization-** Tahazid Tamannur, Sadhan Kumar Das, and Mohammad Meshbahur Rahman

**Writing-original draft-** Tahazid Tamannur, Sadhan Kumar Das, Arifatun Nesa, Foijun Nahar, Nadia Nowshin, Tasnim Haque Binty, Shafiul Azam Shakil, Shuvojit Kumar Kundu, Md Abu Bakkar Siddik, Shafkat Mahmud Rafsun, Umme Habiba, Zaki Farhana, Hafiza Sultana, Anton Abdulbasah Kamil, Mohammad Meshbahur Rahman

**Writing -review and editing-**Tahazid Tamannur, Sadhan Kumar Das, Arifatun Nesa, Foijun Nahar, Nadia Nowshin, Tasnim Haque Binty, Shafiul Azam Shakil, Shuvojit Kumar Kundu, Md Abu Bakkar Siddik, Shafkat Mahmud Rafsun, Umme Habiba, Zaki Farhana, Hafiza Sultana, Anton Abdulbasah Kamil, Mohammad Meshbahur Rahman

## Data availability statement

The datasets generated and/or analyzed during the current study are available from the corresponding author on reasonable request to.

## Notes

### Competing Interest Statement

The authors have declared no competing interest.

### Funding Statement

This study did not receive any funding.

### Author Declarations

This study was approved by the institutional review board of the National Institute of Preventive and Social Medicine (NIPSOM), Bangladesh (Ref No: NIPSOM/IRB/2017/09). Both written and verbal consent were taken before initiating the interview. A brief on the aims and objectives was given to the participants. Participants who agreed were finally included in the study. old: a cross-sectional study in Dhaka

