## Supplemental Table S1-S2 for "Mothers’ knowledge and practices towards oral hygiene of their children aged 5-9 years old: a cross-sectional study in Dhaka": Supplementary file.docx

Table S1. Individual distribution of mothers’ knowledge regarding their children’s oral hygiene

| **Knowledge related variables** | **Incorrect response**  **f (%)** | **Correct response**  **f (%)** |
| --- | --- | --- |
| Importance of brushing teeth | 19(4.7) | 381(95.3) |
| Recommended frequency of tooth brushing | 71(17.7) | 329(82.3) |
| Appropriate time of cleaning teeth | 77(19.2) | 323(80.8) |
| Appropriate duration of cleaning teeth | 282(70.5) | 118(29.5) |
| Different types of toothpaste available | 76(19.0) | 324(81.0) |
| Importance of tongue cleaning | 35(8.7) | 365(91.3) |
| Gingival disease is the most common cause of gum bleeding | 114(28.5) | 286(71.5) |
| Tooth brushing and flossing protect against bleeding gum | 157(39.2) | 243(60.8) |
| Meaning of teeth plaque | 38(9.5) | 362(90.5) |
| Bacteria transmission from mother to child if same utensils are used | 86(21.5) | 314(78.5) |
| Sugary diet causes dental caries | 13(3.2) | 387(96.8) |
| Soft drink causes dental caries | 105(26.2) | 295(73.8) |
| Tooth brushing protects against dental caries | 20(5.0) | 380(95.0) |
| Fluoride toothpaste protects against caries | 246(61.5) | 154(38.5) |
| Dental health affects general health | 36(9.0) | 364(91.0) |

Table S2. Individual distribution of mothers’ practice regarding their children’s oral hygiene

| **Practice related variables** | **Incorrect response**  **f (%)** | **Correct response**  **f (%)** |
| --- | --- | --- |
| Regularity of tooth brushing | 19(4.7) | 381(95.3) |
| Frequency of brushing teeth | 223(55.7) | 177(44.3) |
| Duration of brushing teeth | 281(70.2) | 119(29.8) |
| Aids used for teeth cleaning | 4(1.0) | 396(99.0) |
| Brushing method used | 190(47.5) | 210(52.5) |
| Duration of changing tooth brush | 152(38.0) | 248(62.0) |
| Use of toothpaste | 9(2.2) | 391(97.8) |
| Using fluoride toothpaste | 360(90.0) | 40(10.0) |
| Use of dental floss | 389(97.2) | 11(2.8) |
| Do tongue cleaning | 232(58.0) | 168(42.0) |
| Rinse mouth after eating/drinking | 89(22.2) | 311(77.8) |
| Time of giving sugary food items | 350(87.5) | 50(12.5) |
| Taking to dentist | 399(99.7) | 1(0.3) |
